# OpenSAFELY: Do adults prescribed non-steroidal anti-inflammatory drugs have an increased risk of death from COVID-19?

**DOI:** 10.1101/2020.08.12.20171405

**Authors:** The OpenSAFELY Collaborative, Angel YS Wong, Brian MacKenna, Caroline E Morton, Anna Schultze, Alex J Walker, Krishnan Bhaskaran, Jeremy P Brown, Christopher T Rentsch, Elizabeth Williamson, Henry Drysdale, Richard Croker, Seb Bacon, William Hulme, Chris Bates, Helen J Curtis, Amir Mehrkar, David Evans, Peter Inglesby, Jonathan Cockburn, Helen I McDonald, Laurie Tomlinson, Rohini Mathur, Kevin Wing, Harriet Forbes, John Parry, Frank Hester, Sam Harper, Stephen JW Evans, Liam Smeeth, Ian J Douglas, Ben Goldacre

## Abstract

**Importance:** There has been speculation that non-steroidal anti-inflammatory drugs (NSAIDs) may negatively affect coronavirus disease 2019 (COVID-19) outcomes, yet clinical evidence is limited.

**Objective:** To assess the association between NSAID use and deaths from COVID-19 using OpenSAFELY, a secure analytical platform.

**Design:** Two cohort studies (1^st^ March-14^th^ June 2020).

**Setting:** Working on behalf of NHS England, we used routine clinical data from >17 million patients in England linked to death data from the Office for National Statistics.

**Participants:** Study 1: General population (people with an NSAID prescription in the last three years). Study 2: people with rheumatoid arthritis/osteoarthritis.

**Exposures:** Current NSAID prescription within the 4 months before 1^st^ March 2020.

**Main Outcome and Measure:** We used Cox regression to estimate hazard ratios (HRs) for COVID-19 related death in people currently prescribed NSAIDs, compared with those not currently prescribed NSAIDs, adjusting for age, sex, comorbidities and other medications.

**Results:** In Study 1, we included 535,519 current NSAID users and 1,924,095 non-users in the general population. The crude HR for current use was 1.25 (95% CI, 1.07–1.46), versus non-use. We observed no evidence of difference in risk of COVID-19 related death associated with current use (HR, 0.95, 95% CI, 0.80–1.13) in the fully adjusted model.

In Study 2, we included 1,711,052 people with rheumatoid arthritis/osteoarthritis, of whom 175,631 (10%) were current NSAID users. The crude HR for current use was 0.43 (95% CI, 0.36–0.52), versus non-use. In the fully adjusted model, we observed a lower risk of COVID-19 related death (HR, 0.78, 95% CI, 0.65–0.94) associated with current use of NSAID versus non-use.

**Conclusion and Relevance:** We found no evidence of a harmful effect of NSAIDs on COVID-19 related deaths. Risks of COVID-19 do not need to influence decisions about therapeutic use of NSAIDs.

## Background

The coronavirus disease 2019 (COVID-19), caused by the novel severe acute respiratory syndrome coronavirus 2 (SARS-Cov-2), has been diagnosed in approximately 18 million patients with >690,000 deaths in more than 200 countries as of 5^th^ August 2020.^1^ While most infected people have mild symptoms, several studies reported that people aged ≥60, or those with cardiovascular disease, hypertension, diabetes, chronic respiratory disease and cancer are more likely to have poorer disease prognosis, leading to death.^2–4^

Non-steroidal anti-inflammatory drugs (NSAIDs) are widely prescribed for relief of pain and inflammation in patients with a wide variety of conditions such as acute migraine, osteoarthritis (OA) and rheumatoid arthritis (RA). In the last 12 months, nearly 11 million NSAID prescriptions were dispensed from GP prescriptions in England.^5^ Additionally, some NSAIDs (ibuprofen, aspirin and (in one indication) naproxen) are available over the counter without a prescription from pharmacies and other settings such as supermarkets, with a single brand of ibuprofen alone having sales of approximately £100 million per annum.^6^ Despite their widespread use and tolerable safety profile, non-interventional studies have suggested that NSAIDs may be associated with increased risk of complications of lower respiratory tract infections.^7–15^ These led to a debate over whether NSAIDs would similarly worsen the prognosis of COVID-19. Whilst most studies have focused on the potential for NSAIDs to worsen outcomes in patients with respiratory infections, many complex biological pathways, which are not identical in their actions, are affected by NSAIDs. Indeed, there is also evidence that indomethacin may have protective antiviral effects.^16^

On 14^th^ March, it was recommended in France that patients should avoid the use of NSAIDs due to an apparent worsening of COVID-19 in those taking NSAIDs, based on unpublished reports.^17^ This gained worldwide attention following tweets by the French health minister, including substantial media coverage in the UK^18^ and resulted in the NHS England medical director issuing a directive that paracetamol should be used in preference to NSAIDs^17^ for symptoms of COVID-19. Subsequent reviews by US Food and Drug Administration,^19^ UK Medicines and Healthcare products Regulatory Agency,^20^ and European Medicines Agency^21^ recommended that individuals who currently use NSAIDs for the management of chronic diseases should continue the treatment based on current available evidence in other areas whilst calling for more evidence specifically in patients with COVID-19.

We therefore set out to investigate the association between the use of NSAIDs and deaths from COVID-19 using linked data from over 17 million patients in England.

## Methods

### Study design

We conducted two cohort studies using primary care electronic health record (EHR) data linked to death data from the Office for National Statistics (ONS) between 1^st^ March 2020 and 14^th^ June 2020.

### Data Source

Primary care records managed by the GP software provider The Phoenix Partnership (TPP) were linked to ONS death data through OpenSAFELY, a data analytics platform created by our team on behalf of NHS England to address urgent COVID-19 research questions (https://opensafely.org).^22^ OpenSAFELY provides a secure software interface allowing the analysis of pseudonymized primary care patient records from England in near real-time within the EHR vendor’s highly secure data centre, avoiding the need for large volumes of potentially disclosive pseudonymized patient data to be transferred off-site. This, in addition to other technical and organisational controls, minimizes any risk of re-identification. Similarly pseudonymized datasets from other data providers are securely provided to the EHR vendor and linked to the primary care data. The dataset analyzed within OpenSAFELY is based on 24 million people currently registered with GP surgeries using TPP SystmOne software. It includes pseudonymized data such as coded diagnoses, medications and physiological parameters. No free text data are included.

### Study Populations

We identified two cohorts, anticipating that underlying factors influencing NSAID use and therefore potential biases would differ between them: 1) General Population - All people with at least one prescription for an oral NSAID within the 3 years before study start date (1^st^ March 2020); 2) OA/RA Population - All people with a diagnosis of RA or OA on their primary care record before study start date.

In both cohorts, people with missing data for gender, index of multiple deprivation (IMD) or less than 1 year of primary care records were excluded. We restricted both cohorts to people aged between 18 and 110 years. Aspirin is also used at lower doses in the UK as an antiplatelet to prevent cardiovascular disease,^23^ indicating aspirin users constitute a different population from other NSAID users. We therefore excluded people with a record of an aspirin prescription in the 10 years prior to study start date or a record of either stroke or myocardial infarction ever in primary care before study start date. We also excluded people with a record of gastrointestinal bleeding before study start date, as it is a contraindication to NSAID use. We further excluded people with an asthma diagnosis in the 3 years before study start and a prescription for a short-acting beta agonist (SABA) inhaler within 4 months before study start date as NSAIDs are not recommended in people with asthma due to the risk of bronchospasm.^24^

### Exposures

In the main analysis, we defined current NSAID users as those with any oral NSAID prescription in the 4 months prior to study start, and non-users are those with no record of NSAID prescription in the same time period. Codelists of NSAIDs included are available on <u>codelists.opensafely.org</u> as well as description on how they were derived.

We further examined whether the association varied by types of NSAID, specifically: 1) high/low dose naproxen, 2) COX-2 specific NSAIDs, and 3) ibuprofen. The timeframe used to identify exposure of interest was the same as the main analysis. In the analysis examining doses of naproxen, exposure categories were non-use of NSAID, high dose naproxen (500mg), low dose naproxen (250mg) or any other NSAID based on the strength of the formulation. For COX-2 specific NSAIDs, we split exposure categories as non-use of NSAID, COX-2 specific NSAIDs (defined as celecoxib and etoricoxib) or non-specific NSAIDs. In the analysis examining ibuprofen, exposure categories were non-use of NSAID, ibuprofen or other NSAIDs.

### Outcomes

Follow up for each cohort began on the 1^st^ March 2020 and ended either on date of death or study end date (14^th^ June 2020). If people in the non-use group received a NSAID prescription after 1^st^ March 2020, they were censored at the date of this prescription (eFigure 1).

The outcome was COVID-19 related death as registered in ONS data using ICD-10 codes U07.1 (“COVID-19, virus identified”) and U07.2 (“COVID-19, virus not identified”) listed either as the underlying or any contributing cause of death. The latter ICD-10 code is used when laboratory testing is inconclusive or unavailable.^25^

### Covariates

Potential determinants of exposures and outcomes were identified by reviewing literature and through discussions with practising clinicians. The final list of potential confounders can be seen in Figure 1. Our methodology for creating codelists associated with these confounders has been previously described^22^: this included clinical and epidemiological review and sign-off by at least two authors. Detailed information on every codelist is openly shared at https://codelists.opensafely.org/ for inspection and reuse.

**Figure 1.**
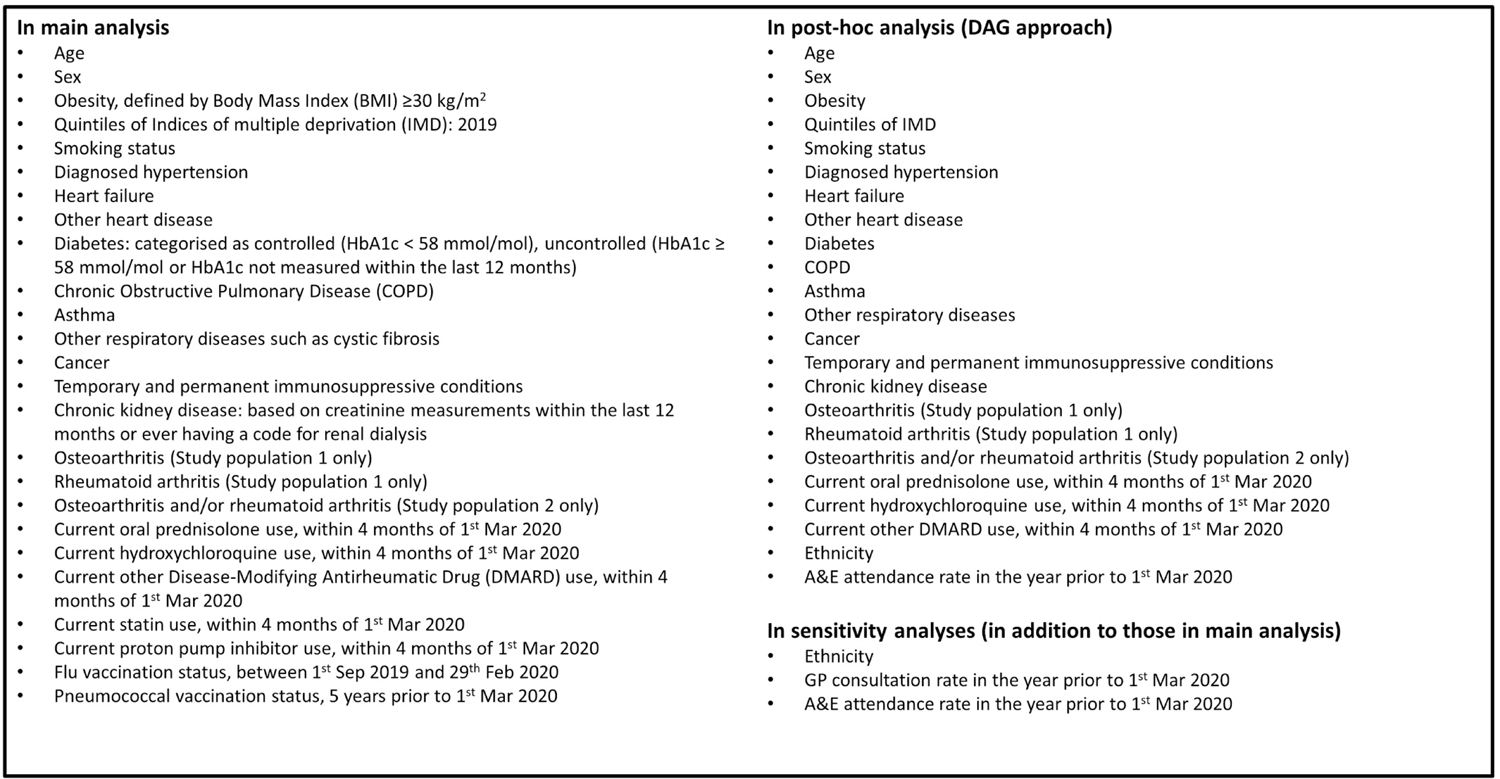
List of covariates Prespecified hypothetical confounders

### Statistical Methods

Baseline characteristics in each cohort were summarized using descriptive statistics, stratified by exposure status. Time to COVID-19 related death was displayed in Kaplan-Meier plots. We additionally present adjusted cumulative mortality curves and the difference between curves using the Royston-Parmar model. We estimated hazard ratios (HR) with 95% confidence intervals for the association between current NSAID use and COVID-19 related death using Cox regression with time since cohort entry as the underlying timescale. We accounted for competing risk by modelling the cause-specific hazard (i.e. censoring non-COVID-19 deaths). We used graphical methods and tests based on Schoenfeld residuals to explore violations of the proportional hazards assumption.

Univariable models, models adjusted for age (using restricted cubic splines) and sex as well as fully adjusted models including covariates listed in Figure 1 were fitted. We stratified the fully adjusted models by geographical regions, defined by Sustainability and Transformation Partnerships (partnerships between NHS organisations and local councils in England^26^), to account for between-region variations. We evaluated the variation by age (under and 70+ years old) and performed likelihood ratio tests to analyse effect modification.

### Quantitative Bias Analysis

We used e-value formulae to calculate the minimum necessary strengths of association between an unmeasured confounder and exposure or outcome, conditional on measured covariates, to fully explain observed non-null adjusted associations (i.e. to move the observed non-null association to the null).^27^

### Sensitivity Analyses

In the main analysis, we did not adjust for ethnicity as it was not anticipated to be a strong confounder and due to a sizable proportion of individuals with missing ethnicity (~23%). Therefore, we additionally adjusted for ethnicity in fully adjusted models and undertook complete case analysis to address missing data. Second, we additionally adjusted for the number of GP consultations and A&E attendance in the past year in fully adjusted models to explore the impact of healthcare seeking behaviours. Third, people with a diabetes diagnosis but not having HbA1c measures in the past year, are likely to have uncontrolled diabetes due to their potential lack of monitoring and management of diabetes. Therefore, we classified these people as uncontrolled diabetes in the main analysis. We tested the robustness of results by separating people with diabetes diagnosis and HbA1c measures ≥58 mmols/mol and those with diabetes diagnosis but without HbA1c measures in the past year into two different categories in the sensitivity analysis. Fourth, we repeated the main analysis with a choice of covariates selected by a Directed Acyclic Graph (DAG) approach (eFigure 2) as a *post-hoc* analysis. Fifth, we varied the definition of currently prescribed an NSAID to within 2 months of 1^st^ March 2020 to assess the sensitivity of exposure definition. Sixth, we repeated the main analysis after excluding indomethacin from all NSAIDs as the exposure of interest, as indomethacin was the only NSAID that was suggested to have antiviral activity against SARS virus.^16^ We also conducted an intention-to-treat analysis by not censoring people who were prescribed NSAIDs after study start date in the non-use group. Finally, we excluded people ever prescribed aspirin before study start date.

### Software and Reproducibility

Data management was performed using Python 3.8 and SQL, with analysis carried out using Stata 16.1. All study analyses were pre-planned unless otherwise stated. All of the code used for data management and analyses is openly shared online for review and re-use (https://github.com/opensafely/nsaids-covid-research). All iterations of the pre-specified study protocol are archived with version control (https://github.com/opensafely/nsaids-covid-research/tree/master/protocol).

### Patient and Public Involvement

Patients were not formally involved in developing this specific study design that was developed rapidly in the context of a global health emergency. We have developed a publicly available website https://opensafely.org/ through which we invite any patient or member of the public to contact us regarding this study or the broader OpenSAFELY project.

## Results

### Main analysis

#### Study population 1: general population

##### Patient characteristics

We included 535,519 current NSAID users and 1,924,095 non-users (Table 1). Median age was 53 years (IQR, 42–64) among current NSAID users and 49 years (IQR, 36–60) among non-users. More women were current NSAID users (59.2%) than non-users (56.7%).

Current NSAID users were more likely to be obese, former smokers, and have a medical history of hypertension, diabetes, other respiratory diseases, cancer, chronic kidney disease, OA and RA than non-users. Current NSAID users were also more likely to have a prescription for statins, proton pump inhibitors, and disease-modifying antirheumatic drugs, and to have had more GP consultations and vaccinations than non-users.

**Table 1.** Demographic and Clinical Characteristics

|  | **Study population 1: General population**<br>**(People prescribed NSAIDs in past 3 years)** |  | **Study population 2: Patients with**<br>**rheumatoid arthritis or osteoarthritis** |  |
| --- | --- | --- | --- | --- |
|  | **Non-use of NSAIDs** | **Current use of NSAIDs** | **Non-use of NSAIDs** | **Current use of NSAIDs** |
| **Total** | **1,924,095** | **535,519** | **1,535,421** | **175,631** |
| **Age as of 1<sup>st</sup> Mar2020** |  |  |  |  |
| 18-<40 | 596,594 (31.0) | 115,478 (21.6) | 32,939 (2.1) | 4,401 (2.5) |
| 40-<50 | 396,273 (20.6) | 102,799 (19.2) | 97,892 (6.4) | 15,808 (9.0) |
| 50-<60 | 423,583 (22.0) | 132,888 (24.8) | 292,583 (19.1) | 45,426 (25.9) |
| 60-<70 | 283,615 (14.7) | 106,169 (19.8) | 417,085 (27.2) | 57,034 (32.5) |
| 70-<80 | 169,290 (8.8) | 62,191 (11.6) | 437,103 (28.5) | 41,387 (23.6) |
| 80+ | 54,740 (2.8) | 15,994 (3.0) | 257,819 (16.8) | 11,575 (6.6) |
| Median, IQR | 49 (36-60) | 53 (42-64) | 68 (58-76) | 63 (55-71) |
| **Sex** |  |  |  |  |
| Female | 1,091,674 (56.7) | 316,844 (59.2) | 952,830 (62.1) | 110,651 (63.0) |
| Male | 832,421 (43.3) | 218,675 (40.8) | 582,591 (37.9) | 64,980 (37.0) |
| **Body mass index** |  |  |  |  |
| <18.5 | 26,253 (1.4) | 5,939 (1.1) | 18,953 (1.2) | 1,219 (0.7) |
| 18.5-24.9 | 483,174 (25.1) | 114,284 (21.3) | 377,596 (24.6) | 31,385 (17.9) |
| 25-29.9 | 575,669 (29.9) | 159,112 (29.7) | 520,008 (33.9) | 55,406 (31.5) |
| 30-34.9 | 331,806 (17.2) | 105,982 (19.8) | 299,254 (19.5) | 40,601 (23.1) |
| 35-39.9 | 137,267 (7.1) | 50,248 (9.4) | 119,714 (7.8) | 20,157 (11.5) |
| 40+ | 70,666 (3.7) | 30,151 (5.6) | 58,615 (3.8) | 12,349 (7.0) |
| Missing | 299,260 (15.6) | 69,803 (13.0) | 141,281 (9.2) | 14,514 (8.3) |
| **Ethnicity** |  |  |  |  |
| White | 1,229,872 (63.9) | 355,452 (66.4) | 1,088,418 (70.9) | 124,381 (70.8) |
| Mixed | 20,415 (1.1) | 4,669 (0.9) | 6,536 (0.4) | 822 (0.5) |
| Asian/Asian British | 151,756 (7.9) | 33,038 (6.2) | 51,832 (3.4) | 7,007 (4.0) |
| Black | 49,486 (2.6) | 10,528 (2.0) | 17,623 (1.1) | 2,094 (1.2) |
| Other | 30,211 (1.6) | 6,926 (1.3) | 10,867 (0.7) | 1,236 (0.7) |
| Missing | 442,355 (23.0) | 124,906 (23.3) | 360,145 (23.5) | 40,091 (22.8) |
| **Index of Multiple Deprivation** |  |  |  |  |
| 1 (least deprived) | 387,308 (20.1) | 107,139 (20.0) | 312,861 (20.4) | 30,687 (17.5) |
| 2 | 385,477 (20.0) | 108,495 (20.3) | 308,559 (20.1) | 32,851 (18.7) |
| 3 | 381,088 (19.8) | 107,220 (20.0) | 307,577 (20.0) | 34,497 (19.6) |
| 4 | 384,665 (20.0) | 106,716 (19.9) | 309,012 (20.1) | 37,427 (21.3) |
| 5 (most deprived) | 385,557 (20.0) | 105,949 (19.8) | 297,412 (19.4) | 40,169 (22.9) |
| **Smoking status** |  |  |  |  |
| Never | 839,206 (43.6) | 219,719 (41.0) | 672,558 (43.8) | 70,249 (40.0) |
| Former | 665,175 (34.6) | 207,375 (38.7) | 694,377 (45.2) | 81,150 (46.2) |
| Current | 388,410 (20.2) | 102,963 (19.2) | 164,760 (10.7) | 23,931 (13.6) |
| Missing | 31,304 (1.6) | 5,462 (1.0) | 3,726 (0.2) | 301 (0.2) |
| **Comorbidities** |  |  |  |  |
| Hypertension | 354,165 (18.4) | 128,108 (23.9) | 626,863 (40.8) | 66,237 (37.7) |
| Heart Failure | 9,477 (0.5) | 2,431 (0.5) | 36,915 (2.4) | 1,412 (0.8) |
| Other Heart Disease | 28,010 (1.5) | 8,731 (1.6) | 58,170 (3.8) | 4,213 (2.4) |
| Diabetes |  |  |  |  |
| Controlled<br>(HbA1c < 58<br>mmols/mol) | 122,377 (6.4) | 41,963 (7.8) | 177,181 (11.5) | 19,469 (11.1) |
| Uncontrolled<br>(HbA1c ≥ 58<br>mmols/mol) | 50,205 (2.6) | 16,474 (3.1) | 58,553 (3.8) | 6,299 (3.6) |
| HbA1c not<br>measured | 4,534 (0.2) | 1,300 (0.2) | 3,675 (0.2) | 417 (0.2) |
| COPD | 42,690 (2.2) | 15,460 (2.9) | 86,193 (5.6) | 8,399 (4.8) |
| Other respiratory<br>diseases | 17,328 (0.9) | 6,197 (1.2) | 38,360 (2.5) | 3,430 (2.0) |
| Cancer | 95,270 (5.0) | 32,144 (6.0) | 174,839 (11.4) | 15,984 (9.1) |
| Immunosuppression | 9,263 (0.5) | 2,890 (0.5) | 8,496 (0.6) | 994 (0.6) |
| Chronic kidney<br>disease | 51,696 (2.7) | 17,568 (3.3) | 165,270 (10.8) | 11,172 (6.4) |
| Osteoarthritis | 368,747 (19.2) | 162,905 (30.4) | 1,476,214 (96.1) | 162,905 (92.8) |
| Rheumatoid arthritis | 28,626 (1.5) | 21,434 (4.0) | 94,851 (6.2) | 21,434 (12.2) |
| **GP consultations** |  |  |  |  |
| Median, IQR | 5 (2-10) | 8 (4-13) | 6 (3-11) | 8 (5-14) |
| Min, Max | 0, 626 | 0, 576 | 0, 585 | 0, 360 |
| **A&E attendance** |  |  |  |  |
| Median, IQR | 0 (0-0) | 0 (0-0) | 0 (0-0) | 0 (0-0) |
| Min, Max | 0, 127 | 0, 153 | 0, 77 | 0, 59 |
| **Vaccination** |  |  |  |  |
| Flu | 434,986 (22.6) | 161,897 (30.2) | 807,032 (52.6) | 86,559 (49.3) |
| Pneumococcal | 116,372 (6.0) | 44,808 (8.4) | 193,115 (12.6) | 24,701 (14.1) |
| **Medications** |  |  |  |  |
| Statin | 223,756 (11.6) | 87,276 (16.3) | 416,818 (27.1) | 47,161 (26.9) |
| Proton pump inhibitors | 268,804 (14.0) | 342,125 (63.9) | 372,180 (24.2) | 137,423 (78.2) |
| Oral prednisolone | 38,988 (2.0) | 16,039 (3.0) | 61,305 (4.0) | 8,237 (4.7) |
| Hydroxychloroquine | 8,076 (0.4) | 6,673 (1.2) | 16,815 (1.1) | 5,104 (2.9) |
| Other DMARDs | 20,742 (1.1) | 16,787 (3.1) | 48,752 (3.2) | 12,699 (7.2) |

Abbreviations: COPD, Chronic obstructive pulmonary disease; DMARDs, Disease-modifying antirheumatic drugs.

##### Univariable and Multivariable Results

eFigures 3 and 4 present time to COVID-19 related death in Kaplan-Meier plots and adjusted cumulative mortality plots respectively. We identified 829 COVID-related deaths in the general population (eTable 1 in the supplement). The crude HR for current NSAID use was 1.25 (95% CI, 1.07–1.46), compared with non-use in the univariable model (Figure 2). After adjustment for age and sex, we observed no evidence of difference in risk of COVID-19 related death associated with current NSAID use (HR, 1.08, 95% CI, 0.93–1.27). Similar results were found in the fully adjusted model (HR, 0.95, 95% CI, 0.80–1.13). There was no evidence suggesting that the HR differed by age in all adjusted models (eTable 2). We did not detect deviations from the proportional hazards assumption (eTable 3, eFigure 5).

**Figure 2.**
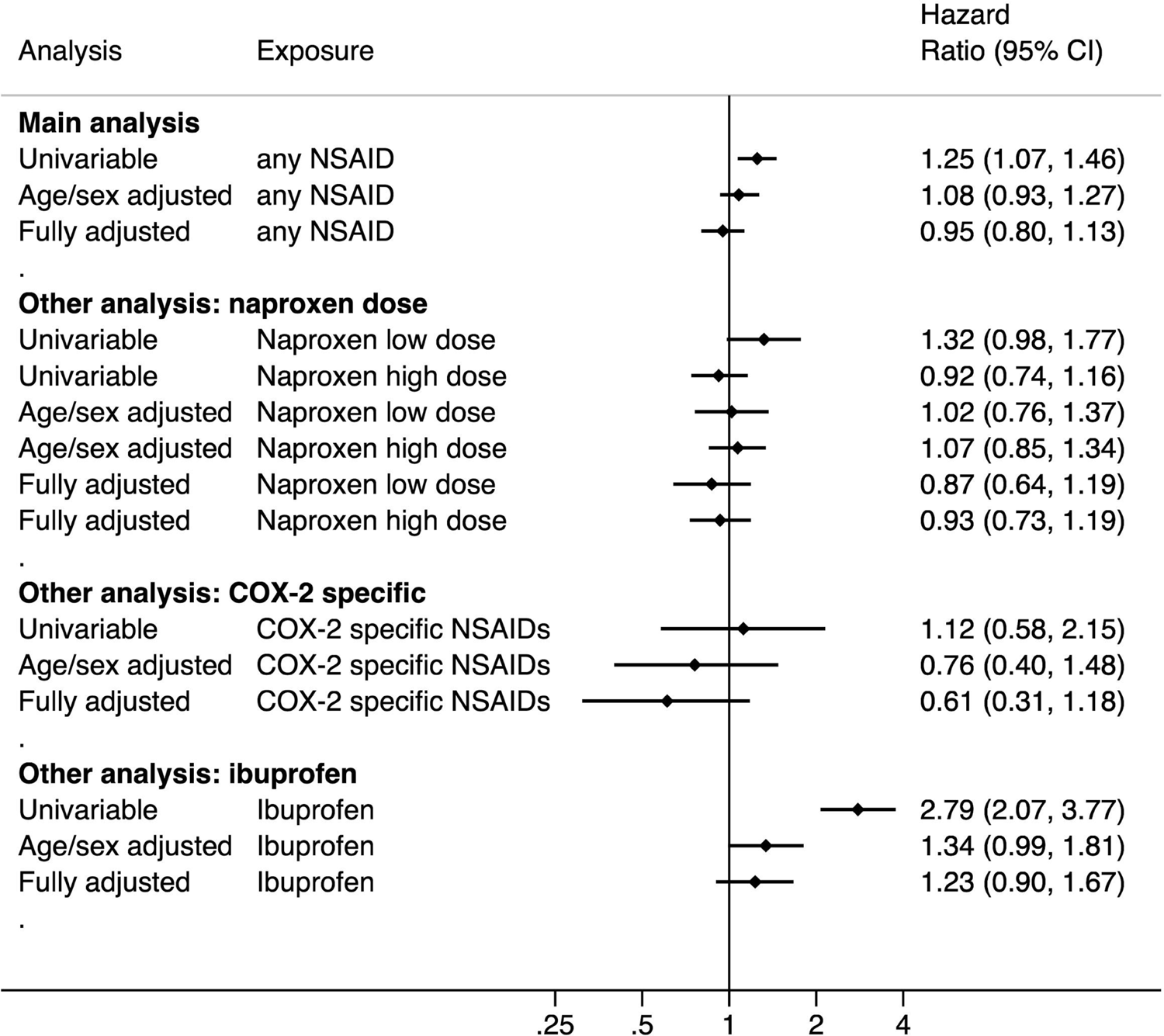
Hazard ratios of the association between current use of NSAIDs and COVID-19 related death in the general population.

#### Study population 2: RA/OA population

##### Patient characteristics

We included 175,631 current NSAID users and 1,535,421 non-users (Table 1). A higher proportion of people aged 70+ were included in this population than the general population. Median age was 63 years (IQR, 55–71) among current NSAID users and 68 years (IQR, 58–76) among non-users. Relative to current NSAID users, non-users were older at study start date. Approximately 60% of individuals were women in both groups.

Current NSAID users were more likely to be obese, more deprived, former/current smokers, and to have had more GP consultations and a prescription for proton pump inhibitors and disease-modifying antirheumatic drugs than non-users. However, non-users were more likely to have comorbidities than current NSAID users.

##### Univariable and Multivariable Results

eFigures 6 and 7 present time to COVID-19 related death in Kaplan-Meier plots and adjusted cumulative mortality curves respectively. We identified 2,564 COVID-related deaths in the RA/OA population (eTable 1). The crude HR for current NSAID use was 0.43 (95% CI, 0.36–0.52), compared with non-use in the univariable model (Figure 3). After adjustment for age and sex, the HR was 0.84, 95% CI, 0.70–1.00). In the fully adjusted model, we observed a lower risk of COVID-19 related death associated with current use of NSAID, compared with non-use (HR, 0.78, 95% CI, 0.65–0.94). *Post-hoc* analyses showed that adjustment for proton pump inhibitors had the largest impact on moving the estimate away from null (eTable 4). Similarly, there was no evidence suggesting that the HR differed by age in all adjusted models. We did not detect deviations from the proportional hazards assumption (eTable 3, eFigure 8).

**Figure 3.**
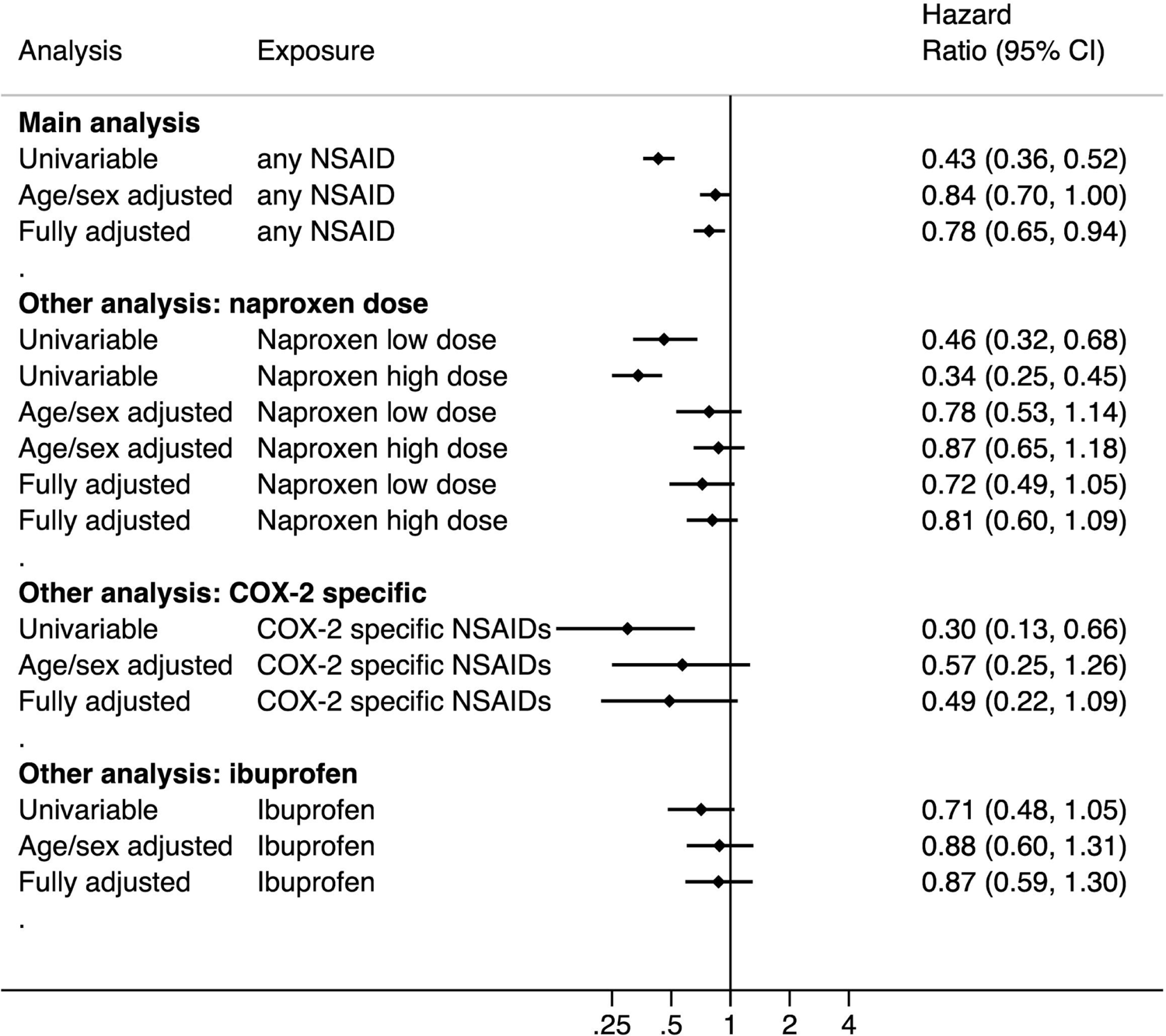
Hazard ratios of the association between current use of NSAIDs and COVID-19 related death in the rheumatoid arthritis or osteoarthritis population.

### Analyses investigating different types of NSAIDs

The baseline characteristics stratified by different types of NSAIDs are presented in eTables 5–10 in the supplement. eFigures 9–10 present time to COVID-19 related deaths by types of NSAIDs in Kaplan-Meier plots.

#### Naproxen dose

In the general population, the fully adjusted HRs were 0.87 (95% CI, 0.64–1.19) for current use of naproxen low dose, 0.93 (95% CI, 0.73-1.19) for current use of naproxen high dose, and 1.01 (95% CI, 0.80-1.29) for current use of other NSAIDs, compared with non-use (Figure 2 and eTable 11). In the RA/OA population, the fully adjusted HRs were 0.72 (95% CI, 0.49-1.05) for current use of naproxen low dose, 0.81 (95% CI, 0.60–1.09) for current use of naproxen high dose, and 0.79 (95% CI, 0.69-1.05) for current use of other NSAIDs, compared with non-use (Figure 3 and eTable 11).

#### COX-2 specific NSAID

In the general population, the fully adjusted HRs were 0.61 (95% CI, 0.31–1.18) for current use of COX-2 specific NSAIDs, and 0.97 (95% CI, 0.82–1.16) for current use of non-specific NSAIDs, compared with non-use (Figure 2 and eTable 12). In the RA/OA population, the fully adjusted HRs were 0.49 (95% CI, 0.22–1.09) for current use of COX-2 specific NSAIDs, and 0.80 (95% CI, 0.66–0.98) for current use of non-specific NSAIDs, compared with non-use (Figure 3 and eTable 12).

#### Ibuprofen

In the general population, the fully adjusted HRs were 1.23 (95% CI, 0.90–1.67) for current ibuprofen use, and 0.89 (95% CI, 0.74–1.07) for other NSAIDs use, compared with non-use (Figure 2 and eTable 13). In the RA/OA population, the fully adjusted HRs were 0.87 (95% CI, 0.59–1.30) for current ibuprofen use, and 0.76 (95% CI, 0.61–0.93) for other NSAIDs use, compared with non-use (Figure 3 and eTable 13).

### Sensitivity analyses

In the sensitivity analysis when we excluded people who were ever prescribed aspirin, we did not observe any difference in risk of COVID-19 related death associated with current use of NSAIDs compared with non-use (fully adjusted HR 0.84, 95% CI, 0.69–1.03) in RA/OA population (eTable 14). In the *post-hoc* analysis when we used a DAG approach to select covariates, we observed no difference in risk of COVID-19 associated with current NSAID use, compared with non-use (HR, 0.86, 95% CI, 0.72–1.03) in the full cohort of patients with RA/OA without adjusting for ethnicity. A marginal decreased risk of COVID-19 was observed in the complete case cohort of patients with RA/OA, without adjusting for ethnicity (HR, 0.80, 95% CI, 0.64-0.99) (eTable 15). The estimate remained the same in the complete case analysis, additionally adjusted for ethnicity. The results of all other sensitivity analyses were broadly similar to those of the main analyses (eTables 16–21).

### Quantitative bias analysis

To fully explain the fully adjusted HR (0.78) or the upper bound of the 95% CI (0.94) in the RA/OA population, an unmeasured confounder would need to be associated (conditional on measured covariates) with either non-use, relative to current NSAID use, or COVID-19 mortality by at least risk ratio (RR) of 1.88 [effect estimate] or 1.29 [upper bound] and with both non-use and COVID-19 mortality by at least RR of 1.28 [effect estimate] or 1.06 [upper bound] (eFigure 11).

## Discussion

### Summary

Based on routinely collected data, our study showed no overall increased risk of COVID-19 related death associated with current NSAID use in adults, compared with non-use. This was consistently seen across all primary, secondary and sensitivity analyses.

We observed a small decreased risk of COVID-19 related death amongst current NSAID users in the RA/OA population but not in the general population. In a *post-hoc* analysis informed by a DAG which captures the complexity of relationships between variables, this protective effect was somewhat attenuated, suggesting it is not a robust finding and is subject to model variable selection. Moreover, our main analysis in the RA/OA population might also be subject to residual confounding as the NSAID users were markedly younger and tended to have fewer comorbidities than those not taking an NSAID. As demonstrated in quantitative bias analysis, an unmeasured confounder of only moderate strength could fully explain this observed association.

### Findings in Context

It was postulated that NSAIDs might delay diagnosis and thus clinical care, by masking the symptoms of a worsening infection.^7,11–13,28^ *In-vivo* and *in-vitro* cellular studies also show that NSAIDs weaken the immune response to pathogens by limiting the local recruitment of innate immune cells and reducing antibody synthesis but the immunomodulatory effects of NSAIDs are not fully understood.^29,30^ Notably, these proposed mechanisms are not specific to COVID-19. Recently, it has also been suggested that ibuprofen upregulates angiotensin-converting enzyme 2 (ACE2)^31^ which has a role in binding SARS-Cov-2 to target cells and could increase the risk of developing severe COVID-19 disease through this route.^32^ In contrast, other animal studies reported that administration of recombinant ACE2 might alleviate lung injury in people with respiratory infection.^33,34^ The evidence is conflicting and it remains unknown whether the findings can be generalised to humans.

In line with our results, three observational studies reported no evidence of a harmful effect of NSAID use on COVID-19 severity among patients with COVID-19^35–37^ but they were of much smaller sample size and not all were general population based, limiting generalisability.^36^ A population-based case-control study which investigated the association between renin-angiotensin-aldosterone system blockers and COVID-19 diagnosis found no association between NSAIDs and COVID-19 diagnosis.^38^ In contrast, a cohort study using data from eight hospitals in the United States reported a lower odds of mortality associated with NSAID use prior to hospitalization among patients with COVID-19 (adjusted odds ratio, 0.56, 95% CI, 0.40–0.82).^39^ However, the patient characteristics, stratified by NSAID exposure and the covariates adjusted for were not clear.

While our study mainly focused on current NSAID use for routine clinical care, there are some ongoing clinical trials investigating the role of NSAIDs in management of COVID-19. They are due to complete later this year or next year, including investigations of whether adding naproxen to standard of care can manage the symptoms of respiratory distress caused by COVID-19 (NCT04325633^40^); whether inhaled or lipid ibuprofen can reduce severity and progression of lung injury in patients with severe acute respiratory syndrome due to COVID-19 (NCT04382768^41^; NCT04334629^42^); and whether a drug cocktail that includes indometacin can improve clinical outcomes in patients with mild COVID-19 (NCT04344457^43^).

### Strengths and weaknesses

The greatest strength of this study was the power we had to examine the association between NSAIDs and COVID-19 death, particularly on types of NSAID as our dataset included medical records from almost 24 million individuals. Our study is further strengthened by the use of two different study populations for comparisons to understand the impact of confounding by indication. The breadth of data available in primary care also allows us to account for a wide range of potential confounders. Additionally, we pre-specified our analysis plan and have openly shared all analytical code. We also recognize possible limitations. First, we do not know whether patients truly took the medications as prescribed. Second, the supply of NSAIDs “over the counter” without a prescription is not captured in GP systems underpinning OpenSAFELY. However, “over the counter” purchases are likely to be for ibuprofen alone, used for acute, irregular conditions such as minor pain or fever, which might affect the results in Study 1 because people prescribed NSAIDs for different indications. However, this is unlikely to impact the result in the RA/OA population as GPs in England are still required to prescribe NSAIDS for long-term conditions such as RA/OA.^44^ A further limitation is that we do not capture all additional medicines commonly used in the treatment of RA. In England, a small number of medicines for long term conditions are supplied routinely by hospitals directly to patients for reasons of reimbursement, safety or administration.^45^ This includes biological treatments such as adalimumab and infliximab and we, along with others, have advocated for the release of this data but access remains restricted.^46,47^ Access to this data is important, as biological treatments might be preferentially prescribed in patients with more comorbidities, resulting in unmeasured confounding in our RA/OA population. It is possible that the higher rates of comorbidities in the non-users of NSAIDs might be because some of those with comorbidities are being given biologics.

### Policy Implications and Interpretation

As our study found no evidence of a harmful association between NSAIDs and COVID-19 death, we recommend that people requiring long-term NSAID treatment continue their treatment as prescribed during the COVID-19 pandemic. OpenSAFELY has delivered rapid insights into the outbreak of COVID-19 including the effect of medicines on the disease.^22,48^ We have demonstrated in this study on NSAIDs that it is feasible to address specific hypotheses about medicines in a transparent manner in response to speculation and calls by regulatory bodies for more evidence. We will use the OpenSAFELY platform to further inform the global response about drug treatments during the COVID-19 emergency.

## Conclusions

We found no evidence of a harmful effect of NSAIDs on COVID-19 related deaths. People currently prescribed NSAIDs for their long-term conditions including rheumatoid arthritis and osteoarthritis should continue their treatment as part of their routine care.

## Data Availability

NHS England is the data controller; TPP is the data processor; and the key researchers on OpenSAFELY are acting on behalf of NHS England. This implementation of OpenSAFELY is hosted within the TPP environment which is accredited to the ISO 27001 information security standard and is NHS IG Toolkit compliant; patient data has been pseudonymised for analysis and linkage using industry standard cryptographic hashing techniques; all pseudonymised datasets transmitted for linkage onto OpenSAFELY are encrypted; access to the platform is via a virtual private network (VPN) connection, restricted to a small group of researchers, their specific machine and IP address; the researchers hold contracts with NHS England and only access the platform to initiate database queries and statistical models.

## Acknowledgements

We are very grateful for all the support received from the TPP Technical Operations team throughout this work; for generous assistance from the information governance and database teams at NHS England / NHSX.

## Information Governance and Ethical approval

NHS England is the data controller; TPP is the data processor; and the key researchers on OpenSAFELY are acting on behalf of NHS England. This implementation of OpenSAFELY is hosted within the TPP environment which is accredited to the ISO 27001 information security standard and is NHS IG Toolkit compliant;^49,50^ patient data has been pseudonymised for analysis and linkage using industry standard cryptographic hashing techniques; all pseudonymised datasets transmitted for linkage onto OpenSAFELY are encrypted; access to the platform is via a virtual private network (VPN) connection, restricted to a small group of researchers, their specific machine and IP address; the researchers hold contracts with NHS England and only access the platform to initiate database queries and statistical models; all database activity is logged; only aggregate statistical outputs leave the platform environment following best practice for anonymisation of results such as statistical disclosure control for low cell counts.^51^ The OpenSAFELY research platform adheres to the data protection principles of the UK Data Protection Act 2018 and the EU General Data Protection Regulation (GDPR) 2016. In March 2020, the Secretary of State for Health and Social Care used powers under the UK Health Service (Control of Patient Information) Regulations 2002 (COPI) to require organisations to process confidential patient information for the purposes of protecting public health, providing healthcare services to the public and monitoring and managing the COVID-19 outbreak and incidents of exposure.^52^ Taken together, these provide the legal bases to link patient datasets on the OpenSAFELY platform. GP practices, from which the primary care data are obtained, are required to share relevant health information to support the public health response to the pandemic, and have been informed of the OpenSAFELY analytics platform. This study was approved by the Health Research Authority (REC reference 20/LO/0651) and by the LSHTM Ethics Board (reference 21863).

## Guarantor

BG

## Contributorship

Contributions are as follows:

Conceptualization LS BG ID;

Data curation CB JP JC SH SB DE PI CM;

Formal Analysis AYSW BM CM JB;

Funding acquisition BG LS;

Information governance AM BG CB JP;

Methodology ID AYSW AS LT KW KB CTR EW SJWE LS JB CM AJW BM SB BG;

Disease category conceptualisation and codelists BM CM AJW RC AS CTR PI SB DE CB JC JP SH HD HC KB SB AM LT ID HM RM HF;

Ethics approval HC EW LS BG;

Project administration AYSW BM CM AS AJW CTR WH CB SB AM LS BG;

Resources BG LS;

Software SB DE PI AJW CM CB FH JC SH;

Supervision ID LS BG;

Visualisation AYSW JB KB;

Writing (original draft) AYSW BM CM ID JB;

Writing (review & editing) AYSW BM CM AS AJW KB JB CTR EW HJC HD SB CB AM DE PI HM LT RH KW HF RC SJWE LS ID BG;

All authors were involved in design and conceptual development and reviewed and approved the final manuscript.

**Abbreviations**
COVID-19: coronavirus disease 2019
NSAIDs: non-steroidal anti-inflammatory drugs
HRs: hazard ratios

